# ‘‘Now, I know my life is not over!’: Introduction and Adaptation of the RESPECT HIV Intervention, OraQuick, and Trauma-Informed Care for Female Victims of Non-Partner Sexual Violence in Haiti

**DOI:** 10.1101/2022.03.11.22272013

**Authors:** Guitele J. Rahill, Manisha Joshi, Berlande Blaise, Cherelle Carrington, Phycien Paul, Caron Zlotnick

**Affiliations:** College of Behavioral & Community Sciences (CBCS), University of South Florida, Tampa, Florida, USA; Department of Behavioral Medicine and Psychiatry, Rockefeller Neuroscience Institute, West Virginia University; OREZON Cité Soleil, Port-au Prince, Haiti; College of Public Health (COPH), University of South Florida, Tampa, Florida, USA; Women & Infants Hospital, Center for Women’s Behavioral Health, Providence, Rhode Island, USA

**Keywords:** RESPECT HIV intervention, Haitian girls and women, victims of non-partner sexual violence, Adaptation of evidence-based interventions, RAPID HIV testing

## Abstract

**Introduction:** In the Cite Soleil (CS) shantytown of Haiti, non-partner sexual violence (NPSV) is widespread, involves multiple assailants who do not use condoms and inflict intentional coital injuries. HIV prevalence in Haiti is 2.2%, CS HIV prevalence is 3.6% shame, guilt, self-blame and societal stigma impede access to HIV testing/treatment in a context of low confidentiality. In that context, NPSV victims often succumb to AIDS. Culturally adapted evidence-based HIV interventions (EBIs) can increase HIV awareness and reduce HIV risk.

**Methods:** Following the ADAPT-ITT model, we used purposive sampling to recruit and interview key stakeholders (age 18 and older) in four focus groups (Victims and health providers, as part of adaptation of an EBI HIV (RESPECT) with an orally administered RAPID HIV antibody test (OraQuick) to increase HIV awareness and testing and to reduce HIV risk for victims of NPSV (N=32, 8/focus group). We also introduced trauma-informed care (TIC) to address the post-assault trauma of NPSV victims. Stakeholders were introduced to RESPECT, participated in RESPECT role plays, interpreted OraQuick HIV screen results after viewing a demonstration of a sample collection, and provided feedback on TIC. ATLAS.ti facilitated thematic content analysis of focus group transcripts.

**Results:** Participants unanimously (100%) reported that RESPECT, OraQuick, and TIC were acceptable, feasible, and useful for increasing HIV awareness, reducing shame, guilt, and trauma, and empowering NPSV victims to reduce the risk of HIV acquisition/transmission in future consensual relationships.

**Conclusion:** Establishing the acceptability, feasibility and effectiveness of RESPECT, OraQuick, and TIC in CS is a crucial first step towards responding to the HIV prevention and trauma needs of NPSV victims.

## Introduction

Worldwide, more than 35 million people live with the human immunodeficiency syndrome (HIV). ^[1-2]^ However, beginning in 2019, crucial funds earmarked for HIV services were diverted to stemming the seemingly relentless assault of COVID-19. ^[1]^ Indeed, since the inception of COVID-19, progress toward the 90/90/90 target of ending HIV in the world has been disrupted. ^[2]^ Consequently, nearly 700,000 additional persons died from HIV-associated complications, and over 1.5 million new HIV infections were documented. ^[2]^ HIV testing and care rates only resumed a positive trajectory in late 2020. ^[2]^ Without targeted engagement of vulnerable persons in the HIV continuum of care (prevention, diagnosis, treatment and care of persons living with HIV [PLHIV]), newly established global 95/95/95 targets will not be achieved. ^[2]^

HIV testing and engagement in treatment and care are crucial for neglected communities in resource-constrained settings such as Haiti, where vulnerable communities still lack even basic HIV knowledge. ^[3]^ HIV knowledge and risk reduction counseling are fundamental to increasing HIV testing rates among high-risk individuals who do not or cannot access HIV testing. ^[4]^ Knowing HIV status can empower individuals with knowledge that informs action to protect themselves and others. ^[1-2; 4-6]^ Biomedical interventions such as pre-exposure prophylaxes (PrEP and PeP) have been powerful resources in preventing HIV, but recent research indicates that they are not sufficient to explain HIV risk reduction in high-risk individuals, nor can the effect of evidence-based behavioral interventions be minimized or discounted. ^[5]^ Thus, behavioral interventions remain important in the fight to eradicate HIV and are crucial to help researchers and other key stakeholders increase HIV testing in resource-constrained settings. ^[6]^ Behavioral interventions are especially needed to increase the accessibility of acceptable HIV services for key populations who also experience violence that may increase HIV risk. ^[2;6]^

Key populations, groups whose susceptibility to HIV is high, account for more than 66% of new HIV infections around the world. ^[2]^ Key populations currently include “men who have sex with men, pregnant women, transgender individuals, people who are sex workers, people who inject drugs, and people in prison and other closed settings”. ^[2]^ Subgroups of key populations in resource-constrained settings are possibly at heightened risk for HIV, given that funding local ministries of health does not necessarily translate into investment of HIV awareness, testing, and treatment resources in their underserved communities.

A recent study identified heightened HIV susceptibility of Haitian women who survive particularly brutal sexual violence (NPSV) in Haiti, locally called *kadejak*. ^[11]^ *Kadejak* involves multiple aggressors, deliberate reproductive injuries, psychological trauma, and heightened risks of sexually transmitted infections (STIs). ^[7-11]^ Addressing the HIV susceptibility of women who survive NPSV in that context must take into account the characteristics of the NPSV, that their HIV risk profile involves sexual risk behaviors that are forced and beyond their control, and that they have little awareness of the risks associated with *kadejak* experiences. ^[3; 11-13]^ Evidence-based interventions for HIV (EBI HIV) can increase these women’s HIV awareness, lead to HIV testing, and reduce HIV risk. There is no current contextually derived EBI HIV being implemented in Haiti. Culturally adapted EBI HIV can increase Haitian NPSV survivors’ knowledge of how HIV is transmitted, encourage HIV testing, place crucial HIV interventions within their control, and reduce the risk of HIV acquisition/transmission in post-NPSV consensual relationships.

### Purpose of the Study

The present study had five objectives: (1) to obtain additional data regarding the characteristics of NPSV described in previous studies; (2) to introduce the RESPECT EBI HIV^[14]^ to Haitian women who self-identify as victims of *kadejak*/ NPSV, who based on their rejection of the term “survivor” and on their self-description as *viktim*,^[11]^ obligate our use of victims to refer to them in this work; (3) to obtain key stakeholders’ (victims, peers, and local health providers) first-hand narratives regarding the RESPECT EBI, its delivery method, and its pertinence to their NPSV experiences. (This third objective is consistent with the WHO’s recommendation that members of key populations and their peers be involved in determining pertinent services and modes of service delivery. ^[1-2]^ It is also consistent with previous requests by CS victims, that:

> …*kadejak* is our problem, so any training that would help us must include our voices. Even in speaking with you today, a heavy load has been taken off my back; so, help us to work; do some training for us—for us to take leadership of ourselves and with other victims like us also—just so that we could improve a little. This is the best work that could take place in [CS]. We thirst for it! We are waiting for it until the day that we die. ^[15, p. 19]^

The fourth objective was to obtain participants’ feedback regarding a self-administered oral HIV antibody test (OraQuick)^[16]^ and to assess their ability to correctly interpret its results. The fifth and final objective was to introduce trauma-informed care (TIC)^[16]^ and to obtain participants’ feedback regarding TIC as an approach for helping victims. We introduced TIC because victims in previous studies endorsed symptoms of PTSD as a result of their NPSV experiences, and women from the same neighborhood who had experienced NPSV in addition to neighborhood violence had higher rates of trauma than nonvictims. ^[7-8]^ Victims in previous studies also reported shame and discomfort experienced in health care settings among reasons that they did not seek post-NPSV care, so that even if those settings could provide PeP, they were not accessing it. ^[7-8; 11]^

### Study Setting: Haiti

Haiti, located in the Caribbean Sea, provides a unique setting to introduce an EBI HIV for NPSV victims. Haiti’s HIV prevalence is approximately 2.2%, a tremendous reduction from over 12% in the past three decades. ^[18]^ This progress toward eradication of HIV in Haiti is due in part to bringing testing, treatment, and other services to members of key populations and to battling racist and ethnocentric accusations of Haitians as carriers of HIV and AIDS. ^[18-21]^ Despite these triumphs, Haitian women have consistently borne a heavier HIV risk and burden than men. ^22-24^ In 2019, HIV prevalence for Haitian women aged 15-49 was 2.3%, compared to 1.5% for men, over 86,000 women were living with HIV compared to 62,000 men, and 2700 of 5700 new HIV cases involved girls/women aged 15-49 years compared to 2300 men. Additionally, HIV knowledge regarding how to prevent contracting the virus was less than 40% among men and women in the 15-24-year-old range, suggesting that HIV prevention messages may not be reaching them. ^[23]^ Additionally, less than 50% of women aged 15-49 reported condom use at last high-risk sex,^23^ compared to over 70% men, indicating that in addition to low HIV knowledge, women may not feel empowered to negotiate for safer sex. Disparities in HIV knowledge, incidence, and prevalence have been considered the possible “feminization of HIV” in Haiti but may be based on a tendency for Haitian women to seek health services more frequently than men. ^[24]^ Nevertheless, HIV prevalence and knowledge of HIV status among 15- to 49-year-old girls/women fluctuate with their region/neighborhood of residence in Haiti. ^[23-25]^ Additionally, Haiti does not have universal testing and treatment, nor does it have universal antiretroviral therapy (ART) for persons living with HIV, making HIV control difficult. ^[26]^

Haiti’s disproportionate burden of HIV is sustained by several factors. Its position in the Caribbean Sea heightens its susceptibility to annual hurricanes, which, in addition to the notorious 2010 earthquake, weaken its health infrastructure. ^[27]^ Additionally, since Haiti’s independence on January 1, 1804, colonialism, racism, colorism, and Negrophobia (fear/dislike of black people/culture) have justified the ascription of inferiority to blacks and thus the unequal distribution of power/resources to its darkest citizens. ^[20]^ Further, Haiti has endured centuries of repressive leadership and human rights violations, which, combined with the legacy of colonialism, have eroded its natural resources and threatened to overwhelm socioeconomic and human capital. ^[21]^ Thus, with more than 6,000,000 people earning just over US$2 per day and an increasing poverty rate in the last 10 years, Haiti remains the poorest nation in the Latin America/Caribbean region. ^[27]^ Despite its challenges, Haiti’s strengths include the intelligence and resilience of its people, Haitians’ giving nature, and vulnerable communities’ resolute struggles against injustice. ^[28-29]^

#### Haiti’s Cité Soleil

Haiti’s Cité Soleil zone is a densely populated urban shantytown along the Bay of Port-au-Prince and near the Delmas River that holds approximately 350,000 individuals within less than 8 square miles.^30^ Cité Soleil residents endure poor sanitation, lack of potable water, food insecurity, poor police protection, and recurrent exposure to gang violence, either as witnesses or as victims. ^[30-31]^ The zone has been described as the most disadvantaged zone of urban Haiti, with an average daily household income that is nearly half the national average. ^[30]^ Given Haiti’s history of colonialism, racism, and colorism, it is not surprising that nearly 100% of Cité Soleil residents are Black. Consequently, there appears to be an unspoken consensus among Cité Soleil residents that Whites are superior and merit socioeconomic dominance and that the darkest or blackest persons are inferior and can be systematically neglected, used, or cast-off. ^[32]^ Cité Soleil residents are currently underrepresented in health research, but nearly three decades ago, being a woman in Cité Soleil between the ages of 20 and 29 years and having more than one sexual partner in the previous year were associated with heightened HIV risk. ^[33]^

Despite the weak infrastructure and bleak socioeconomic context, Cité Soleil has brilliant, industrious people and numerous gifted leaders. ^[29; 34-35]^ Among these are colleagues at OREZON Cité Soleil (French acronym for Organization for the Renovation and Education of the Cité Soleil Zone)^[35]^and local victims and health providers who have long called for HIV prevention and services for survivors of NPSV. ^[11-12; 32]^

#### NPSV in Cité Soleil

As mentioned earlier, NPSV in Cité Soleil is *kadejak*, a term that will be used interchangeably with NPSV in this study when participants are being quoted. ^[11]^ NPSV against women, girls, and transwomen in Haiti’s socioeconomically marginalized neighborhoods occurs most frequently during local and national elections and in the aftermath of disasters, fueled by damaging gender norms that marginalize women and the use of alcohol and drugs by perpetrators. ^[33; 36]^ A typical Cité Soleil Victim is a young woman between 13 and 35 years who is beaten and raped by numerous violent and misogynistic perpetrators. ^[11]^ A previous study indicated that between 50-72% of women in Cité Soleil have endured *kadejak*, as described herein, but new data are needed. ^[11]^ Surveillance data in Cité Soleil are generally scarce, partly because community health agents and human rights advocates avoid the zone due to gang violence in the neighborhood. ^[11; 30]^ However, physicians and community health workers at a local hospital recently indicated that HIV prevalence in its constituents was 3.6%,^[35]^ greater than the national 2.2%. Lacking precise surveillance data from Cité Soleil, a conservative estimate is that 50% of females are between 15 and 49 years old (n≈87,550). Using the lower estimate of 50%, approximately 43,750 girls and women may have experienced NPSV. Given the individual, environmental, societal, and health risks known about victims, adapting an EBI for victims in Cité Soleil is an ethical and moral obligation and a prudent, cost-effective approach relative to a randomized control trial of an EBI that had not been adapted. ^[13]^

### Theoretical framework

Social Justice Theory is at the heart of this study. ^[38]^ Applied to the present study, social justice indicates that even if Victims are not guaranteed civil and human rights of body autonomy and are not recipients of legal remedies for the violent acts of NPSV they endure, in that larger context of extreme poverty, food insecurity, and neighborhood violence, they should at least be equipped with the knowledge of how HIV is transmitted, how to self-screen for HIV antibodies, how to prevent contracting HIV in consensual sexual relationships post-NPSV, and how to reduce risk of transmitting HIV to sexual partners and offspring. The RESPECT EBI could teach and address those issues. Social justice also dictates that victims lacking access to PeP should have access to trauma-informed HIV services that maximize their biopsychosocial safety and security, especially given the nature of the sexual violence they endure.

### The RESPECT EBI

The original RESPECT EBI (hereafter, RESPECT) targeted HIV-negative adults whose risk profiles comprised consensual unprotected sex with multiple partners;^[39]^ it incorporates HIV knowledge, HIV risk-reduction counseling, and rapid HIV testing and was proven effective in reducing HIV risk and increasing HIV prevention in its target population. ^[39]^ RESPECT comprises two sessions of one-on-one counseling by a trained HIV counselor using a structured protocol. ^[39]^

In the first 20-minute session of RESPECT, the counselor assesses HIV risk by eliciting information that clarifies client’s risk behaviors (this can include emphasizing lack of HIV knowledge), challenges inconsistencies between client’s beliefs and behaviors, helps the client develop a strategy to reduce HIV risk (in the present case, increase HIV knowledge, reduce post-assault risk), and, with client’s informed consent, administers an HIV test. ^[39]^ In the second 20-minute session post-HIV test, the counselor conveys the test result, helps the client modify the risk-reduction plan and develop a long-term plan, and provides referrals as needed. Referrals can be for confirmatory HIV testing, for diagnosing STIs, which can increase future susceptibility to HIV, or for support in attaining established HIV risk reduction goals. If a client’s HIV screen result is negative, the goal shifts to enhancing awareness of personal risk and providing assistance to develop an attainable HIV risk reduction plan. Alternatively, if the client screens positive, the focus is on reducing their likelihood of transmitting HIV in future consensual relationships. In the case of victims who are pregnant, the goal would also be to prevent HIV transmission to the offspring.

There have been studies in the United States that minimize the effectiveness of RAPID HIV tests with RESPECT. ^[40-42]^ However, the risk profiles of participants in studies that evaluated the RESPECT EBI with a RAPID HIV test did not comprise victims of forcible non-consensual sex with multiple perpetrators or of low HIV knowledge. Moreover, participants in those studies were aware of their behavioural risks for HIV, voluntarily sought accessible confidential HIV testing in trusted facilities, and often did not return for their HIV test results. ^[40-42]^ Cité Soleil Victims lack HIV awareness, knowledge of status, and other prevention/treatment resources. Additionally, in the present study, we presented OraQuick,^[43]^ a self-administered screen for HIV antibodies that is user-friendly and that provides virtually immediate results; these characteristics of an HIV test are important to enhance perceptions of agency and self-efficacy for NPSV victims whose shame and stigma lead them to avoid traditional health care settings.

### Significance

Our study is significant for several reasons. First, it is a moral and ethical imperative^[36]^ to work toward implementation of an EBI such as RESPECT for high-risk NPSV victims in Haiti because they are understudied, have a unique HIV risk profile comprising non-consensual, coercive, and intentionally injurious sex by multiple men, and have low HIV knowledge. Second, consistent with the WHO’s findings that context-specific HIV prevention and testing methods that are spearheaded by familiar community partners increase HIV testing and engagement in treatment,^[1-2; 4]^ these women have been our partners in the adaptation of the RESPECT EBI for several years and have indicated that any intervention that presumes to help them must include their voices. ^[7-8; 11-12]^ Thus, it would be a grave injustice to deny them the EBI that can reduce their HIV risk if they deem the EBI desirable, acceptable, and feasible to implement in their context. Fourth, coercive sex increases the risks of coital injury, sexually transmitted infections (STIs), and psychological trauma among women of color and was found to cause symptoms of PTSD in NPSV victims previously interviewed. ^[9; 12; 45]^ Presumably, the risks of injuries, HIV, and trauma are greater as the number of assailants increases, and trauma increases HIV risk in survivors by impeding their capacity to negotiate for safer sex and to make beneficial health decisions. ^[8; 45]^ Sixth, there is evidence that inadequate scale-up in EBI and other HIV services in countries such as Haiti is among the obstacles to advance mitigating their HIV burden. ^[2]^ Seventh, Haiti continues to have the second highest estimated HIV prevalence in the world. ^[22]^ Eighth, the Sustainable Development Goals include targets of ending the AIDS pandemic, eliminating violence against women and girls, ensuring their access to sexual and reproductive health, ensuring healthy lives and promoting well-being for all persons by 2030. ^[22]^ Finally, context-specific HIV testing methods spearheaded by familiar community partners can contribute to increased HIV testing and engagement in treatment.^1-2^

### ADAPT-ITT

The ADAPT-ITT method of adapting EBIs is also part of this study’s framework. The ADAPT-ITT method, if correctly executed, achieves adaptation of EBI in 8 phases in which key stakeholders are involved. ^[13]^ In Phase 1: Assessment,^[11]^ we determined Victims’ HIV risk profile, established they have low HIV knowledge and ascertained OREZON’s capacity to adapt and deliver an adapted RESPECT. ^[11; 13]^ In Phase 2: Decision-making,^[12-13]^ we made the decision to adopt and adapt RESPECT, and in collaboration with OREZON colleagues, we translated the manual into Haitian *Kreyòl*. ^[12]^ In the present study, we arrived at Phase 3: Administration/Adaptation. ^[13]^ The Administration phase required conducting a modified theater test to determine if the translated RESPECT EBI, OraQuick, and TIC principles would be viewed as acceptable, understandable, feasible, and desirable by victim and health provider stakeholders. These activities aid in the development of a contextually and linguistically adapted version of RESPECT that has the potential to increase the uptake of an adapted EBI HIV in Haiti. ^[13]^ Thus, at the conclusion of this study, we will incorporate stakeholder-relevant outcomes (i.e., outcomes that are relevant to victims, their relatives, local health providers, OREZON administrators, and policymakers) to refine our adaptation of the RESPECT manual.

### Research Questions

Our research questions were as follows: (1) Are the contextual characteristics of NPSV in our present Cité Soleil sample consistent with those obtained in previous studies, and relatedly, do those characteristics heighten NPSV victims’ HIV susceptibility? ; (2) Are the key concepts in a translated Haitian *Kreyòl* version of the RESPECT EBI understandable and acceptable to NPSV victims and health providers? ; relatedly, 3) is the Haitian *Kreyòl* version feasible to implement in Cité Soleil? ; (4) Can Cité Soleil NPSV victims demonstrate a gain in knowledge of how to administer and interpret results of an oral fluid procedure for detecting HIV antibodies (OraQuick)? ; (5) Can Cité Soleil NPSV victims establish the necessary support that is part of the RESPECT intervention? ; and (6) Is a TIC approach desired by Victims and health providers?

## Methods

We conducted this qualitative study in December 2017. The study was approved by the University of South Florida Institutional Review Board and by Haiti’s National Bioethics Committee at the Ministère de la Santé Publique et de la Population (French Acronym for Ministry of Public Health and Population). We collaborated with administrators and staff at a local social service agency, OREZON. We have had long-term engagement and collaboration with OREZON and with the Cité Soleil community since 2010.

### Sampling and recruitment

Together with OREZON colleagues, we used a purposive sampling method (i.e., criterion sampling) to recruit women who self-identified as victims and local health providers. Criterion sampling is useful when the researcher is engaging key stakeholders that meet some preestablished criteria that are central to the studied phenomena,^[46-47]^ in the present case, identification/experience as NPSV victim or health provider.

Although our sample size was constrained by financial limitations, a sample size such as ours is acceptable when the intent is to access key stakeholders who can provide information-rich data regarding the phenomena being studied rather than generalizing findings from the study. ^[46]^ Additionally, the Administration phase of ADAPT-ITT requires a small sample. ^[13]^ Equally important, our OREZON colleagues informed that gathering large numbers of victims in a town hall format would likely pose risks to victims’ safety from perpetrators and their relatives. Thus, we intentionally aimed for a smaller sample and opted to use focus groups as a modified town hall meeting. Focus groups are useful when a researcher seeks extensive discourse among key stakeholders, seeks guidance regarding a particular resource and is interested in clarifying similarities and differences in stakeholders’ perspectives. ^[48]^

We aimed for 8 individuals per focus group, but taking attrition into account, we overrecruited by 10% per Rabiee’s recommendation,^[49]^ recruiting 40 persons initially. Of these 40, 8 were unable to present for various reasons. To ensure maximum variation in health provider perspectives and inclusion of women’s perspectives regarding RESPECT, OraQuick, and TIC, we intentionally included more female health providers than males. Our final sample size comprised 32 individuals. Thus, there were 2 focus groups with 8 victims per focus group and 2 focus groups with 8 health providers per focus group.

### Research team

The research team included (1) public health and social work colleagues and translators at OREZON; (2) the primary author (PA), who is fluent in English and Haitian *Kreyòl* and has over three decades of experience providing mental health services to Haitians, is a native Haitian and U.S.-based mid-career researcher in Haitian health, particularly HIV, violence against women, and disasters; (3) mid- to late-career scholars with research experience on violence against women, maternal and child health, and adaptation of EBIs; and (4) a graduate student transcriptionist who is fluent in English and Haitian *Krey*lJ*l*.

### Measures

With input from OREZON, we developed a semi-structured focus group guide in Haitian *Kreyòl*, informed by previous studies and by scientific literature regarding EBI adaptation and on previous research with Victims.

### Procedures

OREZON colleagues translated approved consent forms and other study materials to Haitian *Kreyòl*. The PA reviewed translations and back-translations of all documents and approved the final versions. The PA is also a licensed clinical social worker with over 30 years of practice with Haitians and was prepared to provide support for NPSV victims who might experience distress during the focus groups.

To maximize participant safety and privacy, focus groups were conducted at the El Rancho Hotel in Pétion-Ville, a 20-minute car ride from Cité Soleil. OREZON colleagues facilitated transportation. Each focus group lasted approximately 120 minutes, including two 15-minute breaks. Participants received food and drink but no monetary incentive. We met with the two groups of women first, in the morning. Then, we met with the health providers in the afternoon.

Each participant provided informed consent when they satisfactorily summarized in their own words the study’s purpose and voluntary nature, anticipated benefits and risks, and alternatives to participation. The consent process included obtaining participants’ permission to audio record focus group discussions. OREZON colleagues witnessed participants’ signed consent. The PA clarified that their feedback might be included in publications but that their confidentiality and privacy would be maintained as their comments would be de-identified.

The PA used motivational interviewing (MI) procedures to conduct the focus gorups because participants’ HIV risk profile involved coercive NPSV, and it was important to firmly but gently enhance their awareness of risk during role play of the delivery of RESPECT. MI procedures involve expression of respect, empathy, emphasis of safety and confidentiality, demonstration of reflective listening, identification of inconsistency among values, goals, and behavior, and avoidance of judgment and confrontation. ^[50]^ MI techniques prompt clients to consider and voice justification for behavioral changes, an essential component of the RESPECT intervention. MI clarifies that clients are not alone in their experience. ^[50]^ In the present case, MI helped Victim participants evaluate the pros and cons of acquiring behaviors for addressing NPSV consequences (e.g., learning appropriate responses to HIV/STI risks in consensual relationships). ^[50]^

In each focus group, the PA summarized findings from preliminary studies of NPSV in Cité Soleil, defined EBI, summarized the evidence base for EBI HIV and the rationale for introducing RESPECT for Victims, and introduced RESPECT’s core elements, key characteristics, and timeframe. She emphasized that RESPECT is confidential and delivered one-on-one, privately, by a trained HIV counselor. Next, in each focus group, she demonstrated RESPECT via role play with the help of a volunteer.

Following the RESPECT demonstration, the PA introduced the OraQuick oral fluid test and its relevance as a private and confidential HIV self-test for victims. For purposes of confidentiality, participants were not asked their HIV status or asked to submit to an actual OraQuick test; rather, they viewed two videos. The first video demonstrated how to obtain the oral fluid sample and how to interpret the results. ^[43]^ The second video depicted the process of placing the collected sample in the solution bottle that is part of the kit to the result of a reactive test. ^[44]^ Then, the PA defined trauma, connected trauma with Victims’ statements regarding NPSV experiences in previous studies and in the present study, and then introduced the relevance of TIC principles to Victims’ descriptions of their traumatic experiences. Subsequently, participants discussed knowledge gained and satisfaction with RESPECT and with OraQuick, the likelihood of their accepting and using RESPECT with OraQuick, and the relevance and desirability of TIC for Victims. Participants in each focus group provided verbal feedback on OraQuick’s relevance to an adapted RESPECT and the ease of using it and interpreting the result. The joint participation of male and female health providers in those focus groups maximized variety in those groups and potentially enhanced the quality of those discussions. ^[51]^

## Data management

A bilingual transcriptionist first transcribed focus group discussions from Haitian *Krey*lJ*l* to English. The PA read each transcript while listening to the *Krey*lJ*l* recordings to identify discrepancies; these were resolved between the PA and the transcriptionist. The transcripts were loaded as primary documents into ATLAS.ti 6.2, a software that facilitates analysis of textual data. ATLAS.ti permitted us to view primary documents individually and en masse. ATLAS.ti’s hermeneutic framework facilitated storage, management, extraction, exploration, and comparison of meaningful texts within and across primary documents, as well as retrieval, linkage, and interpretation of textual segments and codes, i.e., it permitted constant comparative analysis. ^[52]^

## Data Analysis

The PA began the coding process by reading each transcript and identifying segments of text that addressed: (1) The contextual characteristics of NPSV in our present sample, and, relatedly, if those characteristics heighten Victims’ HIV susceptibility; (2) Contextual obstacles to Victims’ HIV testing and engagement in care; (3) The understandability and acceptability of the RESPECT EBI’s key concepts translated in Haitian *Kreyòl* to NPSV victims and health providers; (4) The feasibility of implementing the Haitian *Kreyòl* version of the RESPECT EBI in Cité Soleil in a trauma-informed fashion; (5) Any knowledge gain demonstrated by Victims regarding administration and interpretation of the OraQuick oral fluid procedure for detecting HIV antibodies; and 6) Cité Soleil Victims’ potential to obtain the necessary social support that is part of the RESPECT intervention.

### Coding

In the open coding phase, we looked for code groundedness and code density. ^[52]^ In ATLAS.ti, code groundedness refers to code frequency, i.e., how often a code is used or the number of quotations linked to the code. Similarly, code density in ATLAS.ti describes the number of other codes to which a given code is linked. We developed a structured codebook^[53]^ to guide the coding process in ATLAS.ti and to assist us in reaching consensus on the codes and their interpretation. Codes from open coding were grouped into categories. As examples, the code “HIV prevention needs” was both grounded and dense in that it was linked to 68 quotations and was also linked to 19 other codes, including “NPSV*/Kadejak*_HIV risk_Multiple perpetrators”, “Age at time of NPSV experience” “Experience of revictimization”, and “Number of NPSV experiences”. Similarly, the code “Acceptability of RESPECT” was linked to “Use of MI Techniques encouraged openness”, “Privacy and confidentiality”, “Heightening awareness of risky sexual behaviours”, “No idea that NPSV constituted HIV Risk”, and “Feasibility of RESPECT with OraQuick” was linked to “Participants learned from role play”, “Recommendations for how to make RESPECT more contextually relevant”, and “Barriers to HIV prevention”. In axial coding, ATLAS. ti’s “network views” feature enabled exploration of relationships among categories and retrieval of related texts that would serve as participant quotes. ^[52]^

#### Establishing rigor in this study

We took numerous steps to establish rigor in this study^[54]^. The PA, as a Haitian *Krey*lJ*l* speaker, served as the moderator. With the participants’ permission, the PA took copious notes on her laptop as a supplement to the audio recordings and to the transcribed notes; these were entered in ATLAS.ti as comments and memos that expanded interpretation of the transcript data. The PA and the bilingual transcriptionist compared the transcripts with the audio recordings. The use of a structured codebook guided analysis of the transcripts as primary documents in ATLAS.ti. ATLAS.ti’s structured framework enabled analysis of individual transcripts and comparison of each transcript with others and enabled comparison of codes and categories among the four transcripts that were the units of analysis.

Additionally, at the end of each focus group, the PA, as a moderator, reintroduced topics that needed clarification and recorded any confirmation or discrepancy on her laptop. The first two authors debriefed after each focus group, discussing participants’ discussions. We also developed a protocol for transcription that reflected modulation in participants’ voices, using italics or capital letters to denote when participants raised their voices, paused extensively, or whispered. To manage the potential of group think, i.e., individual participants’ perspectives being influenced by other participants’ assertions, the PA carefully observed participants’ reaction to other group members’ statements and encouraged each participant to offer divergent perspectives on the different issues discussed; she also gave participants the phone number at the hotel where she lodged and offered them to call her over the week of her stay in the event they had forgotten something or were too embarrassed to share it during the focus groups. ^[54]^ Only one participant from one of the health providers’ groups called, but it was regarding future research opportunities. The PA and the transcriptionist reached consensus both on the content of the transcripts and on the codes applied at each phase of coding, and the PA and second author reviewed those decisions to consensus.

## Results

### Sociodemographic Characteristics

Women in the two NPSV victims’ focus groups ranged in age from 18-41 years (mean = 24), 20% (n=4) reported being married, and 20% (n=4) reported receiving an income for work (as food vendors). The ages of health providers in the two focus groups ranged from 24-42 years (mean = 27.8); they all (100%) reported being single, and 43.75% (7) reported paid employment. Various professions were represented among the health providers, e.g., social workers, nurses, physicians, and psychologists. Below are the findings based on our research questions. Please note that all names used in this manuscript are pseudonyms.

### Characteristics of NPSV in Cité Soleil

As a reminder, the first objective was to obtain additional data regarding the characteristics of NPSV found in previous studies. Descriptions of NPSV in Cité Soleil were consistent with those provided by Victims and health providers in previous studies, i.e., traumatic, non-consensual, brutal, condomless, coerced sex by groups of men who operate without consent and leave persistent biopsychosocial and spiritual injuries in their wake. Denise, a self-described victim from the first focus group, explained:

> It happened in the middle of the night, after I’d fallen asleep. And there were so many men, I couldn’t even count! The worst part of my *kadejak* experience is that it wasn’t something I agreed to…They BURST through the house and took turns on me; and they were rough and brutal (Long pause). When they left, there was a lot of sperm and I spent the next four months bleeding. It’s like the stench stayed in me for months after-physically, mentally, morally. Denise

Likewise, a participant of the second Victims’ focus group declared:

> There were so many of them and they hurt me so badly that since it happened, I’ve had an infection; and it’s not because I’m negligent with hygiene. I take medication, but it never helps. It goes away, but then comes back. *Mwen santi anndan bouboun mwen cho tankou se yon dife ki limen ladann!* (I feel inside my vagina is constantly burning, as if a fire is raging inside it). It really disturbs me-in my body and in my spirit.
>
> Esthène

Being in the early adolescent or pre-adolescent stage at the time of NPSV experience, lack of previous sexual interaction, and the number of assailants increased the physical and psychological trauma of NPSV. A participant, Joanne entreated listeners in the first focus group to place themselves in her shoes when 10 years prior, as a young adolescent and a virgin, she experienced NPSV by more than a dozen men: “Imagine 15-20 men on a child … who had never had sex! God alone knows how much I suffered!” (Moans). Joanne prompted Madelène’s statement regarding her own experience of NPSV prior to reaching adolescence. Madelène also reintroduced the use of substances such as alcohol by her assailants:

> I was severely injured because I was [less than 13] years old and a virgin when it happened. Some of them were drunk on top of me, smothering me, while others strangled me to keep me from screaming. To this day, I still feel pain inside my lower abdomen because there were four men who exerted a lot of force to enter me.

A community health worker from the second focus group confirmed the relationship between alcohol and drug use and the occurrence of NPSV against women in Cité Soleil:

> We are at great HIV risk in Cité Soleil, especially given substance use. We had a neighbor that was close to our home. We saw 21 young men in the early morning hours smoking marijuana, drinking alcohol and smoking something called *pay* (straw) on the roof of her house. It was around 7 in the morning. Something was telling me to look in on her. I found her in the house. She had fainted, and she was covered with blood and blood was dripping from her.

Paula, from the second focus group, reintroduced a finding from previous studies that a young woman can expect NPSV to occur more than once and that pain and STIs are not the only consequences of NPSV in Victims’ lives:

> I’ve had *kadejak* done to me so many times so far in Cité Soleil that at my age, I’ve even had a [child] already-a child of *kadejak* [who] died four months ago. Since then, *m’varye nan sèvèl mwen* (I’m scattered in my brain). I recently spent four months sick in bed away from people and everything.

In light of the contextual characteristics of NPSVs, risks incurred by victims indicate a need for prompt testing, care, and treatment after such events. However, participants in this study cited several obstacles to HIV testing and treatment. In the first focus group, a victim who had just reached the age of majority endorsed having experienced NPSV twice beginning at age 15; she reported long-term symptoms of STIs but denied awareness of HIV risk until her participation in this study:

> I’ve been sick for a long time. Who knows? Maybe it’s HIV. I never even thought of that because I don’t have a partner right now. Until you came to meet with us, I’d never met anyone who helped us understand our need for HIV prevention and care.

Viviane, a participant in the same group added (and others agreed) that she and others like her avoid HIV testing due to fear that their privacy and confidentiality would be violated:

> We all know that even in what they call confidential testing centers in Cité Soleil, the result of your test determines what room they send you to next; so people sitting down waiting to be tested know if you have HIV or not based on which direction you take after they take your blood.

Her concern shame was echoed by Lovelie in the second victims’ focus group, who stated:

> Victims are ashamed because of how they’re viewed; it’s their secret. Because when you admit it happened to you, it’s a joke on you in the community.

The outcomes of shame and stigma were described by Guilaine, a social worker from the first health providers’ focus group:

> The personal shame and societal stigma are such a burden that it’s not unusual for some victims to become so depressed and hopeless that they drink rat poison or bleach to commit suicide.

Jeniver, a social worker from the second health provider focus group, described:

> In Cité Soleil, not everyone gets a chance to participate in programs that teach people how to protect themselves or how to prevent giving HIV to their partners…because people hired to bring the information are afraid to enter the zone because they fear the types of violence that we live with. Not only that, but, the most vulnerable people, the victims, hide so people don’t castigate them; they suffer more because they suffer alone.

### Acceptability of Haitian Kreyòl RESPECT EBI to Victims and Health Providers

As a reminder, another objective in the present study was to introduce the RESPECT EBI HIV to NPSV victims and to health providers to obtain stakeholders’ feedback regarding the RESPECT EBI, its delivery method, and its pertinence to their NPSV experiences. It would be important to determine if the key concepts in a translated Haitian Kreyòl version of the RESPECT EBI were understandable and acceptable to victims and health providers.

Health providers from both focus groups reflected a clear understanding of the RESPECT EBI, as evidenced by the following statement from a community health worker in the health providers’ focus group, identified in this work by the pseudonym Peterson:

> RESPECT, as I understand it, provides psychological support along with knowledge you give the person about their risk of HIV in general, and it includes having the person take an HIV test so they know if they are negative or positive. Whether they’re positive or negative for HIV, you then reinforce the advice you’re giving them through counseling and motivate them to see reducing HIV and other sexually transmitted disease risks as an important and desirable goal. (PA agrees). You help them plan what behaviors they should have in the future to reduce risk and to prevent infection, right? I think it’s a great intervention. Because if you come and educate me about HIV, I will have a different view of the disease and I will get tested and find out my status. But when I’m not infected, or if I don’t know if I’m infected, then I’m in trouble and the virus will eventually make my system so weak that I can die. In any case, the counseling component will be helpful to help *kadejak* victims *defoule* (unburden) because most of the time when they get raped, they don’t talk; they keep it to themselves. If the person finds themselves in a circle where everyone is sharing, she will find herself wanting to share.

Judeline, a physician from the first health providers’ focus group, indicated that the RESPECT EBI would be important in providing early awareness of risk and referral to testing to victims who often do not receive post-NPSV care until it is too late:

> If time passes after they undergo sexual violence, it’s not good for them, because with time if they have an illness it’s only when it reaches a stage that the symptoms overwhelm then that they will get to a hospital and by then it will already be too late… You find women, even if they have a romantic partner after *kadejak*, they’re still at risk because it’s only when the men are dying, when his symptoms are unbearable that he seeks medical care (Mmhmm) eh okay (laughs)…I see that often. A woman comes and does an HIV test, she positive and the husband refuses to test but insists he’s not positive”. Sometimes I tell the woman, ‘if you’re positive, bring your husband so that he can do the test too’, but women will say, “He does not want to and said that he’s not sick’. So, this RESPECT with OraQuick would put the women in charge of their own health even if the partner refuses to participate.

A social worker from one of the health providers’ focus groups asserted that NPSV victims are reluctant to leave a man who is in a civil union with them due to social norms and economic reasons:

> Most of the cases of HIV infection I come across in my work, the men are the one who infect the women. But, it’s the woman’s existence that is at stake. It’s the man that houses them, gives them everything they need to survive. We have a saying, “*yon koup forme, se yon koup forme*” (A couple formed is a couple formed). The man is the lady’s boss. So if the woman leaves, they will lose all, even though they may fear the man is positive. So, if the man does not want to wear condoms to be intimate with her, she can’t ask him that. So, the RESPECT intervention should be done with men as well as women. We have to change the minds of people about the importance of human rights and certainly the importance of women’s rights!

Viviane, from one of the victims’ focus group demonstrated a clear appreciation of the confidentiality involved in RESPECT:

> What is great about this meeting today and what I like is that you keep reminding everyone that what’s said in here stays here. None of us would want to hear, ‘Oh, I heard such and such about her’ after we leave here (Participants agree in background). That’s right!

Below, Figure 1 illustrates a segment of text from the RESPECT role play, in which Joanne voluntarily participated.

**Figure 1:**
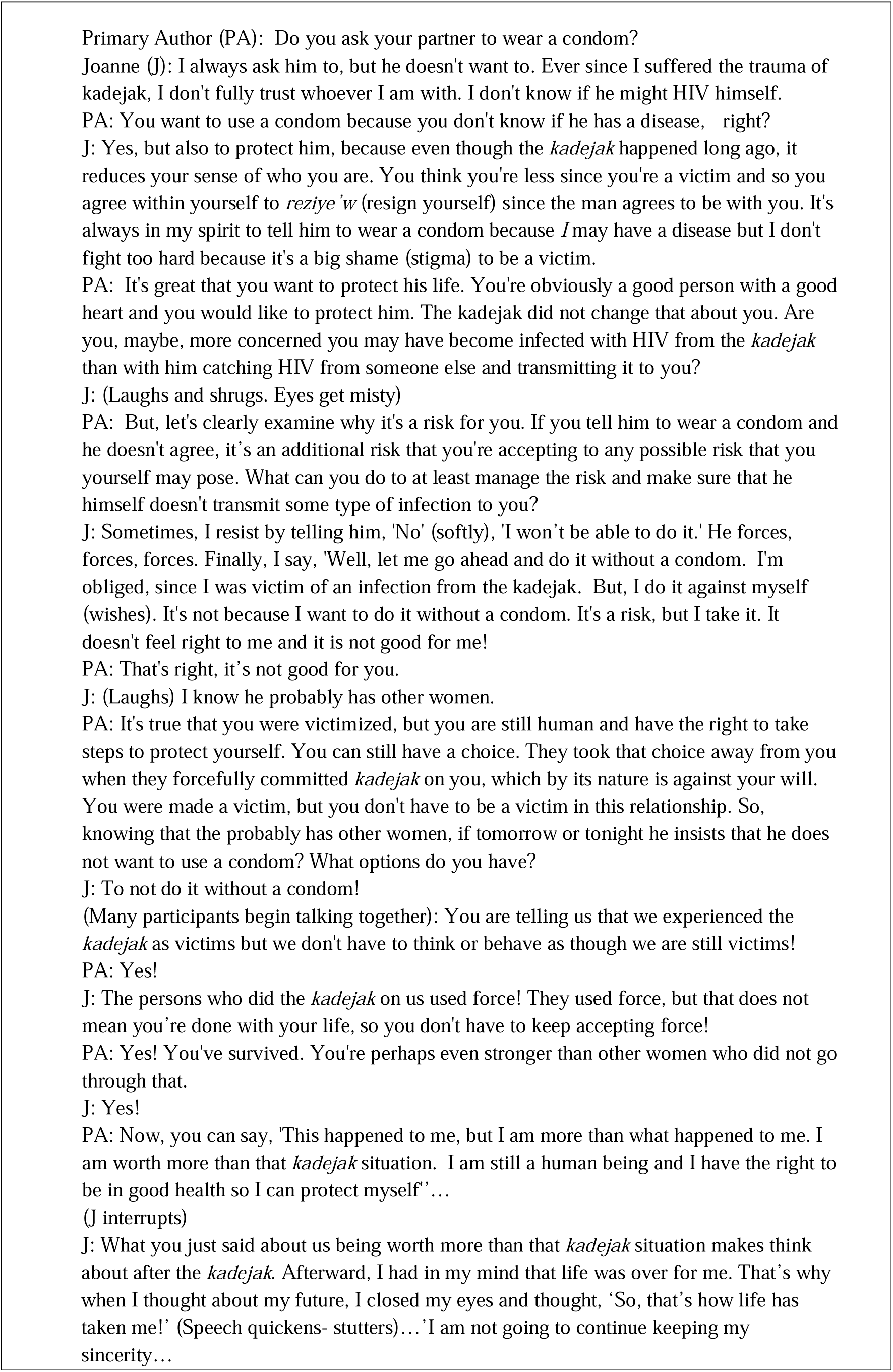
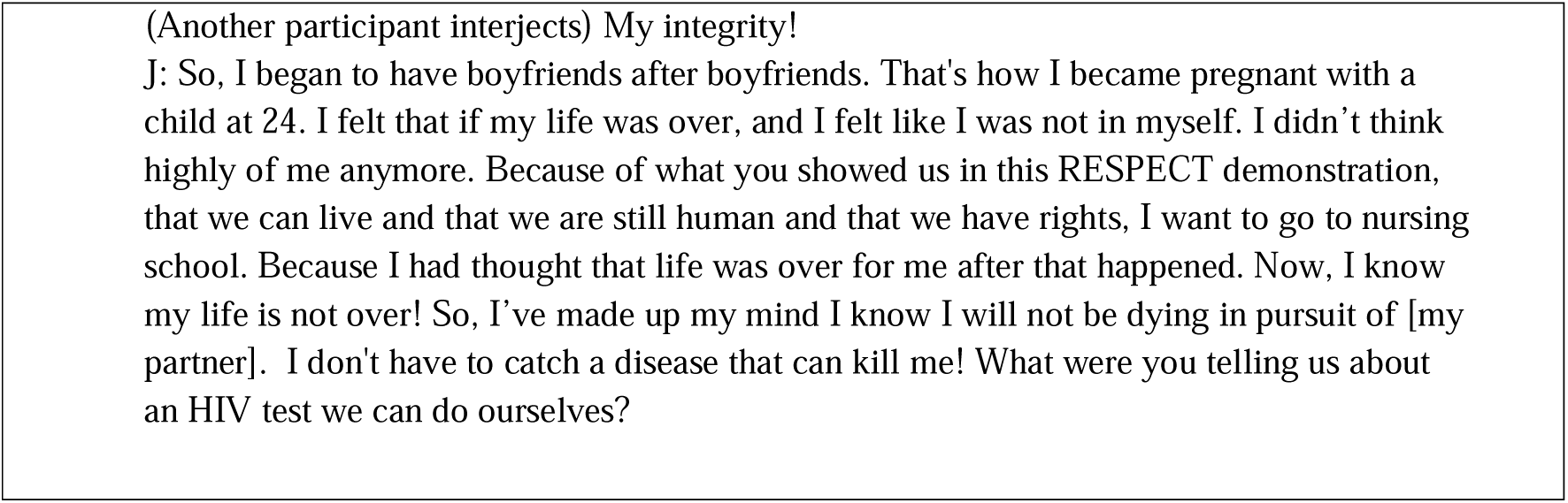
Illustrative Quote for Acceptability, Feasibility and Preliminary Effectiveness of RESPECT for NPSV Victims.

The text in Figure 1 illustrates the complex relationships among NPSV, trauma, shame, lack of awareness that NPSV constitutes an HIV risk, and Victims’ perception that they do not have the right to insist on safer sex practices or to refuse sexual advances in post-NPSV consensual relationships. Figure 1 illustrates that Joanne’s awareness of contracting HIV or some other STI in her current relationship was increased, her misconceptions of herself as damaged goods with no future were gently but firmly challenged, and she became aware of her right to negotiate for safer sex in her consensual relationship with her partner. It also reveals the potential for RESPECT to have an impact beyond the individual receiving the intervention and to foster support for recipients of the intervention.

After facilitating role plays of the RESPECT EBI, in each focus group, the PA solicited participants’ feedback regarding a self-administered oral HIV antibody test, OraQuick, and assessed their ability to correctly interpret its results.

### Participants’ Feedback on OraQuick

All participants in this study successfully interpreted the results of an OraQuick test from a video. Victims, in particular, demonstrated great investment in ensuring that OraQuick is successful in their milieu, offering the best venues for safe storage of the self-test kits and most timely points post-assault to share it with NPSV Victims. Nevertheless, Victims, most of whom were unemployed and whose highest potential daily salary would not suffice to purchase a kit, noted that it would need to be secured to prevent theft. As one of the youngest participants stated, with others concurring emphatically,

> The gang members would steal it and sell it or use it for their own people. You would have to distribute the OraQuick outside of Cité Soleil. That way we could get it and either use it where you give it out, or bring it home with us. This means that the whole RESPECT intervention would have to take place outside of the Cité. We could do it here where we are meeting because it is private and safe.

Stevenson, a psychologist who participated in the health providers focus group also exclaimed at the price of OraQuick and of the potential of gang members to seize it:

> Wow, that OraQuick is so expensive! We could not afford to have it here for Victims despite its value. On the contrary, if word got out that we had such an expensive product, bandits might break in and steal it, or worse yet, it might put our lives at risk. The women from your group are right-this is something that is best done outside of the Cité. The risks are too great; this doesn’t mean it can’t benefit our community members; just we can’t keep it here safely. The gang members would steal it and sell it or use it for their own people.

### Participants’ Feedback on TIC

The fifth and final objective was to introduce trauma-informed care (TIC) to participants as a valuable approach for engaging and supporting NPSV victims. Health providers and women in the victims’ group both agreed that a trauma-informed approach would counteract or mitigate the social stigma and blame encountered by NPSV Victims in Cité Soleil. Janina, a NPSV victim who first experienced NPSV at age 17 and twice since, emphasized the following:

> I only wish I had received that kind of reception after that first time I experienced *kadejak* at least. They treat us like we invited this malediction ourselves. That type of (trauma-informed) care is needed not just for the victim of *kadejak* but also for the whole community. (Several participants speak together, assenting). We just don’t have that here, this feeling for us as victims. If someone is a victim of *kadejak*, rather than blame the victim based on the way they walk or something, I think (trauma-informed care) would have made me receptive to looking for help the second or third time. I didn’t. I can still see the judgment in the doctor’s and nurse’s eyes after that first kadejak. I just stayed home and cried and hoped I wasn’t pregnant since there were so many men who did that to me. Yes, bring that kind of treatment because I am still traumatized—even from the first instance!

The health providers were also receptive to TIC and expressed confidence that they could convince their administrators to adopt TIC as an approach to working not only with NPSV victims but also with all residents of Cité Soleil engaged in health seeking. Stephanie, a nurse participant offered:

> You know, it’s not that you don’t want to show you care for these girls when and if they show up for care after a *kadejak*. It’s that you, as a woman, you are afraid to show too much sympathy lest you be accused of being *sitirèz* (enabling). So you toughen yourself and barely look in their eyes and you thank God it’s not you or your family that this happened to. If you trained the clinics and hospitals in the entire capital about this (TIC), it would be good. Because you never know what burdens the sick person in front of you is carrying in addition to the fever or pain they talk about!

## Discussion

The World Health Organization (WHO) currently aims to reduce new HIV infections in girls and women to less than 100,000,^[56-57]^ engaging PLHIV in counseling and care to reduce the virus to a non-detectable load and reduce its transmission and increase self-testing in highly impacted nations and communities. ^[56]^

In the present study, we sought additional data regarding the characteristics of NPSV found in previous studies and assessed the acceptability and feasibility of implementing a Haitian *Kreyòl* version of the RESPECT EBI together with OraQuick, an oral-fluid HIV antibody screen, using TIC as an approach.

Victims in this study confirmed experiencing non-consensual, unexpected, and intentionally brutal acts of rape, typically by several men, and corroborated previous findings that some girls/women experienced NPSV more than once in their lifetime. Presumably, with each new experience of NPSV, HIV risk and HIV prevention and psychological counseling needs increase. Victims’ age at the time of their initial NPSV experiences emerged as salient; in this small purposive sample, the majority of the Victims were either in late adolescence, were emerging adults, or were young women of childbearing age. Moreover, reports of being virgins at the first NPSV experience further expose them to serious coital injury and entry of sexually transmitted diseases that may be present in the semen of multiple perpetrators. Repeated experiences of *kadejak* by multiple assailants during early adolescence and, in many cases, as a virgin have had potent, adverse effects on Victims in this study, exposing them to lifelong trauma and life-threatening HIV. Moreover, they are further victimized by an inadequate and unempathetic healthcare system—healthcare workers who fail to adequately care for them physically and emotionally once under their jurisdiction, by not protecting the confidentiality of their HIV status after testing, and by blaming them for inviting these salacious atrocities. The “sociocultural milieu that tolerates, excuses, and normalizes sexual assault makes clear labeling a challenge for survivors” ^[56, p. 7]^. Further, “failure to acknowledge the reality of trauma and abuse in the lives of children and the long-term impact this can have on the lives of adults, is one of the most significant clinical and moral deficits of current mental health approaches”.^[57, p. v]^ Ostensibly, this may manifest as complex trauma that can result in a loss of emotional self-regulation, the ability to relate interpersonally, and impairment in one’s mental and behavioral capacity ^[58-59]^. These factors suggest that an adapted RESPECT would need to be trauma informed.

Additional factors suggesting that NPSV victims in Cité Soleil are at heightened risk for HIV include contextual obstacles such as lack of access to primary HIV intervention and confidential HIV testing. Moreover, community stigma leading to guilt, shame, and self-stigma appears to hinder Victims’ ability to negotiate for safer sex even in consensual relationships post NPSV. However, after the introduction of the RESPECT EBI, the self-reported victims said that although they had been victimized, they were not victims. Therefore, instead of living with shame and the stigma of experiencing a kadejak, if these women were exposed to RESPECT EBI, this information could potentially *help* influence the way Victims see themselves and in turn influence how they conduct themselves in future relationships. Furthermore, healthcare workers stressed the importance of providing RESPECT EBI to women at an early age because of its focus on awareness of HIV risk and referral to HIV testing, as NPSV victims often do not receive care after their experience of kadejak until it is too late. Reportedly, even workers who are paid to take this information to the zone refuse to go because of fear for their own lives, so the women of Cité Soleil are not getting the information they need. Health providers in the present study, especially women, may experience moral injury knowing they are being paid to do a specific job to help this community and they are not complying and fulfilling their moral and ethical responsibility. Health providers in the present study potentially experience moral injury recognizing they have an ethical responsibility to care for patients who have experienced *kadejak* and yet, out of fear of being accused as being *sitirèz* (enabling), they “toughen up and barely look into their eyes”. Therefore, if heath care workers could also be exposed to RESPECT EBI, this information could potentially help influence the way they view patients and their moral and ethical role in patient treatment and in turn influence how they are able to care for future patients.

The introduction and role play of RESPECT and of OraQuick were successful in heightening awareness of risk and reflected the potential to empower Victims’ sense of agency and self-advocacy, as, for the first time, they became aware of their right to negotiate for safer sex in consensual relationships. Despite low formal education, Victim participants’ responses to the RESPECT and OraQuick presentations suggest that an adapted RESPECT could contribute to post-traumatic growth in Victims, inspire hope, i.e., “Now, I know my life is not over,” equip them to negotiate for safer sex in consensual relationships, and provide them with a self-administered HIV test that they can interpret. Having hope restored highlights the importance of RESPECT EBI and the relevance of this study. Therefore, researchers and health care workers need to focus not only on the incidence and prevalence of trauma, but also on Victim’s capacity for resilience, hope, and post-traumatic growth after these harrowing and horrifying experiences of *kadejak*.

### Limitations

This study has several limitations. First, its findings may not be generalizable to socioeconomically non-equivalent neighbourhoods in Haiti. Nevertheless, the potential impact of an adapted RESPECT with OraQuick extends to Citè Soleil’s adjacent Bel Air, Carrefour, and Martissant zones, all of which face similar socio-structural challenges of poverty, gender-based violence, NPSV, and HIV risk.

A second limitation is in our use of purposive sampling and a relatively small sample. However, Morse justifies purposive sampling procedures and a small sample size in cases where the researchers desire interaction, dialogue, and exchange of ideas among participants with an unfamiliar set of issues and when the sample is sufficiently large to elicit usable and pertinent data from key stakeholders. ^[46]^

## Conclusion

Given Haiti’s disproportionate burden of HIV, it may be useful to begin to view NPSV as a “concentrated subepidemic [in Cité Soleil]…and [it] would, therefore, also be valuable to calculate and report on the indicators that relate to [Victims] as members of a key population at higher risk”. ^[55, p. 23]^

The United Nations and the Pan American Health Organization emphasize the need for Combination HIV prevention, i.e., addressing biopsychosocial, structural, and behavioral factors that relate to HIV incidence and prevalence. Part of HIV prevention for Victims as a high-risk subpopulation of Haitians (since we are not in a position to prevent NPSV in their community) is increasing the uptake and implementation of a targeted EBI HIV that promotes risk awareness and behavior change before and after experiencing NPSV. In elucidating neighbourhood conditions and risks that heighten HIV for NPSV victims in our study, we add to the literature on resource development for psychosocial aspects of HIV prevention, treatment, and service planning. RESPECT with OraQuick, delivered in a trauma-informed manner, can have a tremendous and sustained impact on reducing HIV incidence and on improving the well-being of NPSV victims in Cité Soleil and on reducing the risks of HIV transmission in future consensual relationships.

## Data Availability

All data produced in the present work are contained in the manuscript.

## Notes

### Competing Interest Statement

The authors have declared no competing interest.

### Funding Statement

This study was funded by the University of South Florida HIV Research Group

### Author Declarations

The study was approved by the University of South Florida Institutional Review Board and by the National Bioethics Committee of Haiti.

## References

1. World Health Organization. HIV/AIDS. 2020. www.who.int/news-room/fact-sheets/detail/hiv-aids.

2. World Health Organization. WHO and partners urge countries to fast-track implementation and scale-up of HIV self-testing and other innovative HIV testing approaches in Asia and the Pacific. 2021. www.who.int/news/item/16-03-2021-who-and-partners-urge-countries-to-fast-track-implementation-and-scale-up-of-hiv-self-testing-and-other-innovative-hiv-testing-approaches-in-asia-and-the-pacific.

3. Joshi M, Rahill GJ, Rice C, Paul P, Maio A, Burris C. Back to Basics: Correlates of HIV risk in a community sample of Haiti. MedRxiv, Cold Spring Harbor Laboratory Press; 2020. https://doi.org/10.1101/2020.10.10.20210476

4. Fonner VA, Denison J, Kennedy CE, O’Reilly K, Sweat M. Voluntary counseling and testing (VCT) for changing HIV-related risk behavior in developing countries. The Cochrane Database Syst Rev. 2012;9(9):CD001224. https://doi.org/10.1002/14651858.cd001224.pub4

5. O’Reilly K, Fonner V, Kennedy C, Yeh P, Sweat M. The paradox of HIV prevention: Did biomedical prevention trials show how effective behavioral prevention can be? AIDS. 2020;34(14):2007–11. https://doi.org/10.1097/qad.0000000000002682

6. World Health Organization. World AIDS Day 2020 - WHO Director-General Calls for Renewed Efforts to End AIDS. 2020. www.who.int/multi-media/details/world-aids-day-2020-who-director-general-calls-for-renewed-efforts-to-end-aids

7. Rahill G, Joshi M, Lescano C, Holbert D. Symptoms of PTSD in a sample of female victims of sexual violence in post-earthquake Haiti. J Affect Disord. 2015;173:232–8. https://doi.org/10.1016/j.jad.2014.10.067

8. Joshi M, Rahill G, Rhode S. Comparison of trauma symptoms among nonpartner sexual violence victims and nonvictims in urban Haiti’s Cité Soleil neighborhood. J Black Psychol. First published Feb 21, 2021. https://doi.org/10.1177/0095798421997217

9. Aniekwu NI, Atsenuwa A. Sexual violence and HIV/AIDS in Sub-Saharan Africa: An intimate link. Local Environment: The International Journal of Justice and Sustainability. 2007;12(3):313–24. https://doi.org/10.1080/13549830601098289

10. Cenat JM, Smith K, Morse C, Derivois D. Sexual victimization, PTSD, depression, and social support among women survivors of the 2010 Earthquake in Haiti: A moderated moderation model. Psychol Med. 2019;50(15); 2587–98. https://doi.org/10.1017/s0033291719002757

11. Joshi M, Rahill G, Lescano C, Jean F. Language of sexual violence in Haiti : Perceptions of victims, community-level workers, and health care providers. J Health Care Poor Underserved. 2014;25(4):1623–40. https://doi.org/10.1353/hpu.2014.0172

12. Rahill GJ, Joshi M, Hernandez A. Adapting an evidence-based intervention for HIV to avail access to testing and risk-reduction counseling for female victims of sexual violence in post-earthquake Haiti. AIDS Care. 2016;28(2):250–6. https://doi.org/10.1080/09540121.2015.1071773

13. Wingood GM, DiClemente R. The ADAPT-ITT model: A novel method of adapting evidence-based HIV Interventions. J Acquir Immune Defic Syndr. 2008;47(Suppl1):S40–6. https://doi.org/10.1097/qai.0b013e3181605df1

14. Centers for Disease Control and Prevention. Archived Interventions: Compendium of Evidence-Based Interventions and Best Practices for HIV Prevention: Risk Reduction Chapter. 2020. www.cdc.gov/hiv/research/interventionresearch/compendium/rr/archive.html.

15. Joshi M, Rahill GJ, Lescano C, Jean F. Language of sexual violence in Haiti: Challenges for HIV prevention in international resource-poor settings. Society for Social Work Research 2014 Annual Conference. January 15-19, 2014. https://sswr.confex.com/sswr/2014/webprogram/Paper22058.html

16. OraSure Technologies Inc. OraQuick In-Home HIV Test (Package Insert). OraSure Technologies. 2016. www.orasure.com/docs/pdfs/products/oraquick_advance/OraQuick_ADVANCE_PI-US_EN.pdf

17. Brezing C, Ferrara M, Freudenreich O. The syndemic illness of HIV and trauma: Implications for a trauma-informed model of care. Psychosomatics. 2015;56(2):107–18. https://doi.org/10.1016/j.psym.2014.10.006

18. UNAIDS. Communities at the Centre: Defending Rights, Breaking Barriers, Reaching People with Services. Geneva, Switzerland: Joint United Nations. 2019. www.unaids.org/sites/default/files/media_asset/2019-global-AIDS-update_en.pdf.

19. Antoine LB, Pierre L, Page JB. Exclusion of blood donors by country of origin and discrimination against black foreigners in the USA. AIDS. 1990;4(8):818. https://doi.org/10.1097/00002030-199008000-00018

20. Fanon F, Markmann CL. Black skin, white masks. 1967. Originally published by Editions de Seuil, France, 1952 as Peau Noire, Masques Blanc. First published in the United Kingdom in 1986 by Pluto Press London. Newest edition published 2008.

21. Carlsson G. Communities at the Centre: Defending Rights, Breaking Barriers, Reaching People with HIV Services. UNAIDS. 2019. www.unaids.org/sites/default/files/media_asset/2019-global-AIDS-update_en.pdf

22. United Nations. Sustainable Development Goals: Transforming our World - The 2030 Agenda for Sustainable Development. https://sustainabledevelopment.un.org/topics/sustainabledevelopmentgoals

23. Boulos R, Halsey NA, Holt E, Ruff A, Brutus JR, Quinn TC, et al. HIV-1 in Haitian women 1982-1988. The Cite Soleil/JHU AIDS Project Team. J. Acquir. Immune Defic. Syndr. 1990;3(7):721–8.

24. Ministère de la Santé Publique et de la Population [MSPP] (Haitian Ministry of Health) (2016). Bulletin de Surveillance Epidémiologique contre VIH/SIDA 2016 (2016 Surveillance and Epidemiological Surveillance against HIV/AIDS). 2016;2:1–35. https://mspp.gouv.ht/site/downloads/Bulletin%20de%20Surveillance%20Epidemiologique%20VIH%20Sida%20numero%2012%20MSPP.compressed.pdf

25. Dorjgochoo T, Noel F, Deschamps MM, Theodore H, William D, Wright PF, et al. Risk factors for HIV infection among Haitian adolescents and young adults seeking counseling and testing in Port-au-Prince. J. Acquir. Immune Defic. Syndr. 2009;52(4):498–508. https://dx.doi.org/10.1097%2FQAI.0b013e3181ac12a8

26. Puttkammer N, Parrish C, Desir Y, Hyppolite N, Wagenaar B, Joseph N, et al. Toward universal HIV treatment in Haiti: Time trends in ART retention after expanded ART eligibility in a national cohort from 2011 to 2017. J. Acquir. Immune Defic. Syndr. 2020;84(2):153–61. https://doi.org/10.1097/qai.0000000000002329

27. World Bank. Haiti overview. 2019. https://www.worldbank.org/en/country/haiti/overview

28. Human Rights Watch (HRW). World Report 2019: Haiti. 2019. https://www.hrw.org/world-report/2019/country-chapters/haiti#e81181

29. Haiti Libre. Haiti - FLASH: Results of 9th AF exams, for the West department. Aug 14, 2019. https://www.haitilibre.com/en/news-28489-haiti-flash-results-of-9th-af-exams-for-the-west-department.html

30. World Bank. Social, Urban, Rural and Resilience Global Practice - Latin America and Caribbean Region. Implementation completion and results report on a grant in the amount of XDR 34.4 million to the Republic of Haiti for an Urban Community-driven Development Project (PRODEPUR). 2017. https://documents1.worldbank.org/curated/en/806971509982511950/pdf/ICR-Main-Document-P106699-2017-10-31-19-45-11022017.pdf. Accessed 15 June 2021.

31. Willman A, Marcelin LH. If they could make us disappear, they would! youth and violence in Cité Soleil, Haiti. J Community Psychol. 2010;38(4):515–531. https://doi.org/10.1002/jcop.20379

32. Rahill GJ, Joshi M, Shadowens W. Best intentions are not best practices: Lessons learned while conducting health research with trauma-impacted female victims of nonpartner sexual violence in Haiti. J Black Psychol. 2018;44(7):595–625. https://doi.org/10.1177%2F0095798418810054

33. Duramy BF. The double weakness of girls: Discrimination and sexual violence in Haiti. Stanford J. Int’l. Law. 2008;44(1):147–203. https://digitalcommons.law.ggu.edu/pubs/4/

34. Cousteau P, Knight M. Urban oasis offers hope to Haiti’s poorest. 2013. https://www.cnn.com/2013/07/08/world/americas/urban-oasis-offers-hope-haiti/index.html

35. OREZON. Accueil (Welcome). 2020. https://orezoncs.wordpress.com/

36. Rahill GJ, Joshi M, Galea J, Ollis J. Experiences of sexual and gender minorities in an urban enclave of Haiti: despised, beaten, stoned, stabbed, shot and raped. Cult Health Sex. 2020;22(6):690–704. https://doi.org/10.1080/13691058.2019.1628305

37. Patel V, Farmer P. The moral case for global mental health delivery. Lancet. 2020;395(10218):108–9. https://doi.org/10.1016/s0140-6736(19)33149-6

38. Bell LA. Theoretical foundations for social justice education. In: Adams M, Griffin P, editors. Teaching for diversity and social justice. 1–14. New York: Routledge; 2007. pp.1–14.

39. Department of Health and Human Services [DHHS], Centers for Disease Control and Prevention [CDC]. Effective interventions. https://www.cdc.gov/hiv/effective-interventions/index.html

40. Metcalf C, Douglas J, Malotte C, Cross H, Dillon B, Paul S, et al. Relative efficacy of prevention counselling with rapid and standard HIV testing: A randomized, controlled trial (RESPECT-2). Sex Transm Dis. 2005;32(2):130–8. https://doi.org/10.1097/01.olq.0000151421.97004.c0

41. Johnson CC, Kennedy C, Fonner V, Siegfried N, Figueroa C, Dalal S, et al. Examining the effects of HIV self-testing compared to standard HIV testing services: A systematic review and meta-analysis. J Int AIDS Soc. 2017;20(1);21594. https://doi.org/10.7448/ias.20.1.21594

42. Metsch L.R, Feaster D.J, Gooden L, Schackman B. R., Matheson T, Das M, et al. Effect of risk-reduction counselling with RAPID HIV testing on risk of acquiring sexually transmitted infections: The AWARE randomized clinical trial. JAMA. 2013;310(16): 1701–10.

43. OraQuick RAPID test instruction. 2012. https://www.youtube.com/watch?v=5FBWORY91J4

44. Oraquick Advance HIV 1/2 test. 2013. https://www.youtube.com/watch?v=OY1ast9Wyis

45. Wyatt GE, Myers HF, Williams JK, Kitchen C R, Loeb T, Carmona JV, et al. Does a history of trauma contribute to HIV risk for women of color? AJPH. 2002;92:660–5. https://ajph.aphapublications.org/doi/epub/10.2105/AJPH.92.4.660

46. Morse, JM. Determining sample size. Qual. Health Res. 2000;10(1):3–5. https://doi.org/10.1177/104973200129118183

47. Patton, MQ. Qualitative evaluation and research methods. 2nd ed. Thousand Oaks, CA: Sage; 1990.

48. Krueger RA, Casey MA. Focus Group: A practical guide for research. 5th ed. Thousand Oaks, CA: Sage; 2015.

49. Rabiee F. Focus-group interview and data analysis. Proceedings of the Nutrition Society. 2004;63:655–660. https://doi.org/10.1079/PNS2004399

50. Sobell LC, Sobell MB. Motivational interviewing strategies and techniques: Rationales and examples. 2008. http://www.ncjfcj.org/sites/default/files/MI%20Strategies%20&%20Techniques%20-%20Rationales%20and%20examples.pdf

51. Dahn B, Woldemariam AT, Perry H, Maeda A, Glahn, DV, et al. Strengthening primary health care through community health workers: Investment case and financing recommendations. 2015. https://www.who.int/hrh/news/2015/CHW-Financing-FINAL-July-15-2015.pdf?ua=1. Accessed 15 June 2021.

52. Corbin, J, Strauss, A. Grounded theory research: Procedures, canons, and evaluative criteria. Qual Sociology. 1990;13:3–21. https://link.springer.com/article/10.1007%2FBF00988593

53. Fonteyn ME, Vettese M, Lancaster DR, Bauer-Wu S. Developing a codebook to guide content analysis of expressive writing transcripts. Appl Nurs Res. 2009;21:165–8. https://doi.org/10.1016/j.apnr.2006.08.005

54. Kidd PS, Parshall MD. Getting the focus and the group: Enhancing analytical rigor in focus group research. Qual Health Res. 2000;10(3):293–308. https://doi.org/10.1177/104973200129118453

55. Joint United Nations Programme on HIV/AIDS (UNAIDS). Global AIDS Monitoring 2021: Indicators for monitoring the 2016 Political Declaration on Ending AIDS. 2021. https://www.unaids.org/en/resources/documents/2020/global-aids-monitoring-guidelines

56. Saint Arnault D, Sinko L. Hope and fulfillment after complex trauma: Using mixed methods to understand healing. Front. Psychol. 2019;10:2061. https://doi.org/10.3389/fpsyg.2019.02061

57. Kezelman C, Stavropoulos P. ‘The last frontier’: Practice guidelines for treatment of complex trauma and trauma informed care and service delivery. Blue Knot Foundation, Formerly Adults Surviving Child Abuse (ASCA); 2012. https://www.childabuseroyalcommission.gov.au/sites/default/files/IND.0521.001.0001.pdf

58. Courtois CA, Ford JD. Treating complex traumatic stress disorders: An evidence-based guide. Guilford Press; 2009.

59. Cook A, Spinazzola J, Ford J, Lanktree C, Blaustein M, Cloitre M, et al. Complex trauma in children and adolescents. Psychiatr. Ann. 2017;35(5):390–8.

